## Supplementary figures and images for "Spread of endemic SARS-CoV-2 lineages in Russia"

### Fig. S1

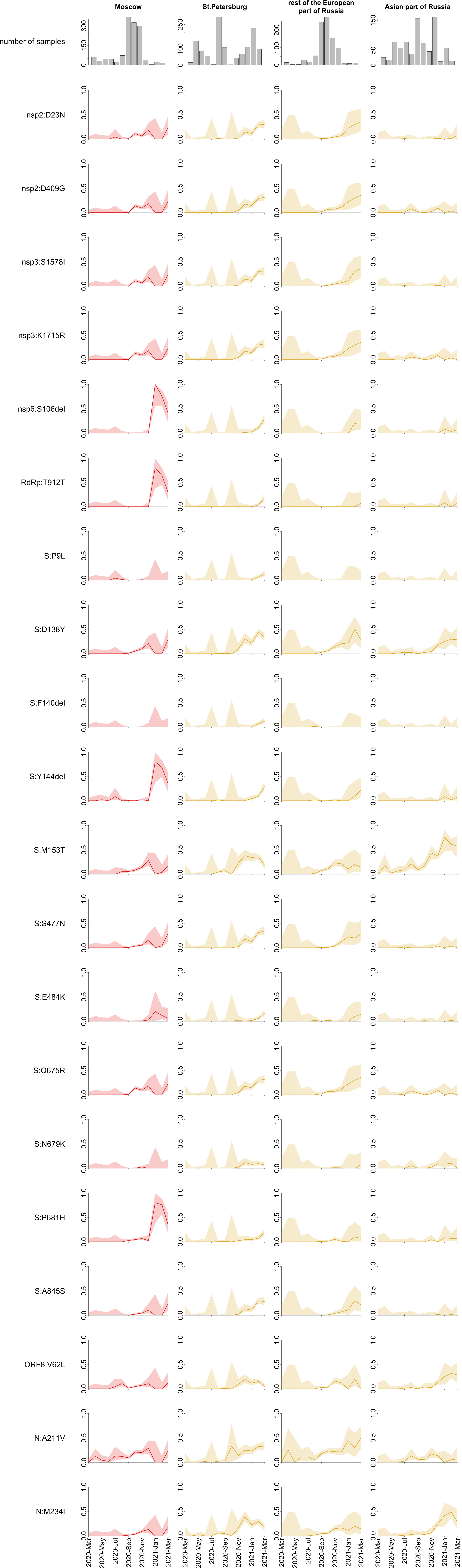
